# Contact tracing and isolation reduces covid-19 incidence in a structured agent-based model

**DOI:** 10.1101/2020.10.06.20207761

**Authors:** Marcus Low, Nathan Geffen

## Abstract

**Background:** The World Health Organization has identified contact tracing and isolation (CTI) as a key strategy to slow transmission of SARS-CoV-2. Structured agent-based models (ABMs) provide a means to investigate the efficacy of such strategies in heterogeneous populations and to explore the impact of factors such as changes in test turnaround times (TaT).

**Methods:** We developed a structured ABM to simulate key SARS-CoV-2 transmission and Covid-19 disease progression dynamics in populations of 10, 000 agents. We ran 10, 000 simulations of each of three scenarios: (1) No CTI with a TaT of two days, (2) CTI with a TaT of two days, and (3) CTI with a TaT of eight days. We conducted a secondary analysis in which TaT values were varied from two to 11. The primary outcome for all analyses was mean total infections.

**Results:** CTI reduced the mean number of infections from 5, 577 to 4, 157 (a relative reduction of 25.5%) when TaT was held steady at two days. CTI with a TaT of eight days resulted in a mean of 5, 163 infections (a relative reduction of 7.4% compared to no CTI and a TaT of two days). In the secondary analysis, every additional day added to the TaT increased the total number of infections – with the greatest increase in infections between four and five days, and the smallest increase between ten and 11 days.

**Conclusions:** In a structured ABM that simulates key dynamics of Covid-19 transmission and disease progression, CTI results in a substantial reduction in the mean number of total infections. The benefit is greater with shorter TaT times, but remained substantial even with TaTs of eight days. The results suggest that CTI may play a critical role in reducing the size of outbreaks and that TaTs should be kept as short as possible in order to maximise this benefit.

## 1 Introduction

Many countries have implemented extended and far-reaching lockdowns in response to the Coronavirus Disease 2019 (Covid-19) pandemic, often at significant economic cost. A key question that has arisen in recent months is what non-pharmaceutical interventions (NPIs) governments can implement in order to exit such lockdowns as safely as possible.[1] Some potential NPIs include promotion of hand-washing and other hygiene measures, promotion of or legal requirement of mask-wearing in public, various physical distancing regulations, border closures, school closures and contact tracing and isolation (CTI).[2]

The World Health Organization (WHO) has endorsed CTI as a key strategy to slow transmission of SARS-CoV-2.[3] CTI has been implemented in a wide range of countries - maybe most notably in China, Singapore and South Korea.[4, 5, 6] One recent Lancet study concluded that “each country should have an effective find, test, trace, isolate, and support system in place”.[1] CTI involves tracing the recent contacts of a person who tested positive for SARS-CoV-2 and isolating these contacts, and in some cases also testing contacts. CTI is traditionally conducted by teams of tracers, but can also be done through, or assisted by various mobile phone applications. Since there are economic, human resource and opportunity costs related to CTI, it is important to understand the effectiveness of such programmes and the factors impacting their effectiveness.

A recent SARS-CoV-2 stochastic transmission modelling study found that ‘minimising testing delay had the largest impact on reducing onward transmissions’ and suggested that, in their model at least, test turnaround times (TaTs) of less than three days were required to get the effective reproductive number under 1.[7] A recent branch process modelling study found that in most scenarios, highly effective contact tracing and case isolation is enough to control a new SARS-CoV-2 outbreak within 3 months, but that ‘the probability of control decreases with long delays from symptom onset to isolation’.[8] Such delays could for example be due to long TaTs.

Structured agent-based models (ABMs) are a way to model the complex consequences of interventions such as CTI on the transmission of SARS-CoV-2. By modelling at the agent level, where each agent represents a person, such models better account for heterogeneity within populations than is the case with compartmental or population-level models that by definition model groups rather than individuals. Though stochastic, neither of the two SARS-CoV-2 models cited above are ABMs. In a recent review, four of seven models investigating CTI for the control of earlier outbreaks of Severe Acute Respiratory Syndrome and/or Middle-Eastern Respiratory Syndrome were agent-based models.[9]

The model we report on here is novel in that it (1) is a high performance ABM calibrated to various Covid-19 parameters, (2) contains structure in the form of households, school classes, workplaces, public transport and neighbourhood blocks, and (3) allows for the modelling of interventions such as CTI and variable TaTs within a structured and heterogeneous population.

### 2 Methodology

We developed a structured ABM, called ABM-Spaces, to simulate key dynamics of the transmission and disease progression of SARS-CoV-2 in a closed population of 10, 000 agents loosely based on the Masiphumelele township in Cape Town South Africa. Each simulation was run for 270 time steps (days). Key elements of the model world are as follows:

#### Agents

Agents are given various attributes, including age, sex, and a variety of variables relating to disease states (exposed, infectious, symptomatic, recovered, dead, etc). Agents are newly created in each simulation, thus introducing variability in age structure, gender balance, and the number of initial infections.

#### Settings

Agents in the model are associated with settings (places). Based on research on tuberculosis transmission in Masiphumelele, the five types of settings in the model are households, school classes, workplaces, public transport (minibus taxis or train carriages) and blocks (collections of households).[10] All agents in the model belong to households. All school-age agents are placed into school classes. A percentage of working-age agents are placed into workplaces. Employed agents are more likely to take taxis. Each instance of the simulation stochastically creates new agents that are associated with newly constituted schools, households and workplaces based on parameters such as employment rate and the average size of school classes.

#### Transmission

Transmission of SARS-CoV-2 in the model occurs not directly from agent to agent, but from agent to setting to agent. On each time step in the model agents are exposed to the transmission risk of the various settings they are associated with. The setting and time step-specific risk of a setting is a product of the number of infectious agents in that setting on that time step and the inherent risk profile of the type of setting. This can be expressed as:

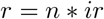

Where *r* is the specific risk faced by a susceptible agent in a specific setting on a specific time step, *n* is the number of positive agents in a setting on a specific time step and *ir* is the inherent risk of the setting (a constant that is different for different types of setting).

For each agent in each setting, on each time step the model draws a random number *k* from a uniform distribution of integers 1 to 10, 000. If *k* is less than *r* transmission takes place. One variation on this is transmission in taxis where transmission only takes place if *k* is less than the square route of *R*.

The parameters for inherent risk (*ir*) associated with different settings were calibrated using two processes: First, setting-related risks were varied so as to reproduce published transmission patterns observed for TB in Masiphumalele.[10] Second, these risks were calibrated so as to produce epidemic simulations with *R*0 values that fall within the range of *R*0 estimates found in the Covid-19 literature.[8]

We calculated *R*0 values using a numerical method that involves deriving *R*0 from the equation *R* = *R*0 \**S/N* – where *S* is the susceptible population and *N* is the total population.[11] Since *R*_*t*_ = 1 at the peak of the epidemic and the fraction susceptible at the peak of the epidemic is known for each simulation, *R*0 can accordingly be solved numerically.

#### Disease progression

On the day an agent becomes infected with SARS-CoV-2, the model draws from a number of distributions and estimates of risk, based on the published Covid-19 literature,[12, 13] to determine (1) whether the agent will become symptomatic and when, (2) when the agent will become infectious, (3) whether the agent will be tested and when, (4) whether the agent will stay home and when, (5) whether the agent will require hospitalisation and when, and (6) whether the agent will die and when. The model remains flexible regarding these values – for example to account for hospitals being full.

#### Testing

The model allows for false negative tests by taking into account published estimates of the variability of test sensitivity based on days since infection. [14] We gave symptomatic agents a likelihood of 0.8 of being tested – although the model can easily be run with other values for this and other variables. The only way for asymptomatic agents to be tested in the model is as part of contact tracing. The model allows for different TaT values to be used. TaT values observed in South Africa ranged from two to greater than 11. [15]

#### CTI

The model can be run with or without CTI. In the default version of the model CTI involves testing 80% of an agent’s household, school class or workplace contacts within two days of the agent testing positive. The test results for these tests become available a default of two days later (TaT equals two) whereupon the same process is repeated for all positive tests – with the modification that agents tested in the last 14 days will not be tested again. In CTI scenarios agents who test positive are isolated from all other agents and cannot further transmit the virus. 80% of agents who are traced, but who do not test positive go into isolation, with the remaining 20% isolating at home. The model can also be run so that a positive agent’s contacts are isolated, but not tested.

Isolation refers to the movement of people with the infection being restricted, voluntarily or legally, while quarantine refers to the movement of people at risk of infection being restricted, voluntarily or legally. The work described here does not differentiate between these distinctions. We therefore use isolation to refer interchangeably to isolation or quarantine.

### 2.1 Analysis

To explore the impact of CTI we held TaT steady at two days and ran 10, 000 simulations without CTI and 10,000 simulations with CTI. We then ran an additional 10,000 simulations with CTI and a TaT of eight days to test whether CTI is still beneficial if there are significant testing delays. In these base CTI scenarios the contacts of everyone who tests positive are tested. Since CTI in the real world sometimes does not involve testing of contacts, we ran an additional 10, 000 simulations of each of the CTI scenarios with the testing of contacts removed.

As a secondary analysis to explore the impact of different testing delays, we ran 10,000 simulations with CTI for each value of TaT from two to 11. This was done with testing of contacts and repeated without testing of contacts.

### 2.2 Programming

Prototype versions of ABM-Spaces were developed in R, but the final version presented here is coded in C++, which is dramatically faster than the R version. The C++ code allows for thousands of simulations to be run in minutes on standard consumer hardware and can easily be modified to explore different research questions in a structured ABM environment. The ABM-Spaces source is available under the GNU General Public License version 3.0. Our code and results are available at https://github.com/nathangeffen/ABM_CTI.

#### 3 Results

CTI reduced the mean number of infections from 5, 577 to 4, 157, a relative reduction of 25.5%, when TaT was held steady at two days. CTI with a TaT of 8 days resulted in a mean number of 5, 163*infections*, a relative reduction of 7.4% compared to no CTI and a TaT of two days. In CTI scenarios where contacts of positive agents were not tested the reduction in the mean number of total infections were more modest. CTI with a TaT of two days and no testing of contacts resulted in a mean of 4, 670 infections, while the same scenario with TaT extended to 8 days resulted in 5, 263 infections (relative reductions of 16.3% and 5.56% respectively compared to no CTI).

In the secondary analysis, longer testing delays were consistently found to lead to increases in mean total infections over 10, 000 simulations. This relationship was not linear. In simulations with testing of contacts increasing TaT from two to three days resulted in an additional 156 infections. This increased to an additional 188 infections when increasing TaT from four to five days (the peak). From there it decreased to 73 additional infections when increasing TaT from ten to 11 days. The difference in mean total infections between simulations with and without testing of contacts was greatest with a TaT of two days and got smaller as TaT increased.

**Table 1:** Results of 10, 000 simulations of each of five scenarios.

| Scenario | Mean total infections | Infections as fraction of no CTI |
| --- | --- | --- |
| No CTI, TaT=2 | 5,577 | 1 |
| CTI, TaT=2 | 4,157 | 0.745 |
| CTI, TaT=8 | 5,163 | 0.926 |
| CTI, TaT=2 (contacts not tested) | 4,670 | 0.837 |
| CTI, TaT=8 (contacts not tested) | 5,263 | 0.944 |

## 4 Conclusions

In a structured ABM that simulates key dynamics of Covid-19 transmission and disease progression, the WHO-recommended strategy of CTI resulted in a substantial reduction in both the mean number of deaths and the mean number of total infections. The benefit is greater with shorter TaT times, but remained substantial even with TaTs of eight days. The difference in mean total infections between TaTs of four and five days is more than double that between nine and ten days. In our model significant numbers of infections can be averted by testing the contacts of persons who test positive – a benefit that is greater with shorter TaTs.

These results suggest that CTI can play a critical role in reducing the size of outbreaks in the real world and that TaTs should be kept as short as possible in order to maximise this benefit. Our findings thus broadly confirm those of Kretzschmar et al and Hellewell et al using a substantially different methodology.[7, 8] Apart from the difference in methodology, our study differs from that of Kretzschmar et al and Hellewell et al in that we asked whether CTI and different TaT values lead to meaningful reductions in total infections, while they explored what level of intervention was needed to reduce the effective reproductive number to below one. It is possible that certain levels of intervention would substantially reduce total infections, but not so much as to bring the effective reproduction number below one.

Since several of our model parameters are highly uncertain, there is uncertainty in the model outputs in addition to the already significant stochasticity in the model. The model outputs should thus at most be considered as illustrative of underlying epidemiological dynamics rather than as an exact prediction of the impact of CTI and TaT in the real world.

The model has several limitations. Many of the Covid-19 epidemiological and disease progression parameters remain highly uncertain. The model does not account for co-morbidities such as diabetes, HIV and TB. Neither does it account for movement in and out of the community, a key element of Covid-19 transmission in the real world. The setting-specific transmission risk parameters in the structured model were deduced from published literature on TB transmission in a Cape Town community (No such South African data yet exists for Covid-19). Finally, we did not attempt to assess the cost-effectiveness of CTI in this study.

## Data Availability

All code and data are available on GitHub.

## Notes

### Competing Interest Statement

The authors have declared no competing interest.

### Funding Statement

No funding received.

